# Impact of Age, Race, and Family History on COVID-19 Related Changes in Breast Cancer Screening among the Boston Mammography Cohort Study

**DOI:** 10.1101/2022.12.20.22283719

**Authors:** Naiyu Chen, David Cheng, Michelle O. Sodipo, Mollie E. Barnard, Natalie C. DuPre, Rulla M. Tamimi, Erica T. Warner

## Abstract

**Background:** We studied women enrolled in the Boston Mammography Cohort Study to investigate whether subgroups defined by age, race, or family history of breast cancer experienced differences in trends of screening or diagnostic imaging rates during the COVID-19 lockdown and had slower rebound in trends of these rates during reopening.

**Methods:** We compared trends of monthly breast cancer screening and diagnostic imaging rates over time between the pre-COVID-19, lockdown, and reopening periods and tested for differences in the monthly trend within the same period by age (<50 vs ≥50), race (White vs non-White), and first-degree family history of breast cancer (yes vs no).

**Results:** Overall, we observed a decline in breast cancer screening and diagnostic imaging rates. The monthly trend of breast cancer screening rates for women age ≥50 was 5% higher (p=0.005) in the pre-COVID-19 period but was 19% lower in the reopening phase than that of women aged<50 (p<0.001). White participants had 36% higher monthly trend of breast cancer diagnostic imaging rates than non-White participants (p=0.018).

**Discussion:** The rebound in screening was lower in women age ≥50 and lower in non-White women for diagnostic imaging. Careful attention must be paid as the COVID-19 recovery continues to ensure equitable resumption of care.

**Funding:** The project was supported by the Breast Cancer Research Foundation (RT). Researchers were supported by the University of Louisville CIEHS P30 ES030283 (NCD), K01CA188075 (ETW), T32CA09001 (NCD, MOS, MEB) P30 ES000002 (JH, FL), and NIH/NCI K00 CA212222 (MEB). This manuscript is the responsibility of the authors and does not represent the official views of the National Institutes of Health.

## Introduction

On February 3, 2020, the United States declared COVID-19 as a public health emergency^1^. Within the next three months, 42 states and territories in the United States issued mandatory lockdown or stay-at-home orders^2^. To reduce transmission and alleviate the burden on the healthcare system, most healthcare facilities chose to decrease capacity and delay non-urgent services, resulting in an almost 90% decline in breast cancer screening immediately after the lockdown orders^3–7^. This disruption in screening then translated into a sharp decrease in the incidence of new breast cancer diagnosis in April 2020 and, as a result, studies have estimated a 7.9% to 9.5% increase in death from breast cancer up to 5 years from diagnosis^7–9^.

The sharp decline in breast cancer screening during the initial lockdown period has been well documented^3,4,10,11^. It is also important to document and understand potential demographic differences in the impact of the pandemic on breast cancer screening to ensure that the COVID-19 pandemic does not exacerbate or create new health disparities in breast cancer early detection. During the pandemic, patients must balance risks of potential COVID-19 infection and other burdens with benefits of screening for other disease types^12^. Older patients and historically marginalized populations face a higher risk of hospitalization or death due to COVID-19^13^.

Additionally, these populations may also be more disproportionately impacted by the pandemic, where they are more likely to experience job loss and a lack of health insurance, and/or to care for other family members due to COVID-19. These are all potential factors that may make them less likely to seek non-urgent preventive care^14–16^. Conversely, individuals with a first-degree family history of breast cancer may place greater priority on breast cancer screening, even during the pandemic^13^.

While multiple studies have described changes in breast cancer screening and diagnosis with the advent of the pandemic, the rate of return to care during the reopening period among different demographic populations has not been fully described^11,17–19^. Building upon this existing literature, this study examines monthly rates of breast cancer screening from before the pandemic through the recovery period following the initial lockdown by age, race, and family history. As the pandemic progresses, it becomes more relevant to shift our focus from the initial impact of the lockdown to the rate of recovery, especially among groups that are already experiencing cancer health disparities prior to the pandemic. This focus on recovery allows researchers to identify potential points of intervention to mitigate and counter existing health disparities.

To better understand the impact of the COVID-19 pandemic on breast cancer screening and diagnostic imaging, we studied participants from the Boston Mammography Cohort Study (BMCS) to investigate whether subgroups defined by age, race, or first-degree family history of breast cancer: 1) experienced differences in trends of screening or diagnostic imaging rates during the COVID-19 lockdown; and 2) had slower rebound in trends of these rates during reopening.

## METHODS

### Study Design and Study Population

In this cohort study, we examined utilization of breast cancer screening and diagnostic imagining among participants in BMCS^20^. BMCS includes women who were enrolled prior to a routine screening mammogram visit within the Mass General Brigham healthcare system (MGB) between 2006-2014 (N=2,821). Participants completed a survey at the time of enrollment, which collected information on demographic characteristics, including date of birth and self-reported race as well as breast cancer risk factors including first-degree family history of breast cancer.

We used cancer registry and electronic health record data to identify and exclude patients with a personal history of breast cancer (in-situ or invasive) at the start of the study period, leaving 2,392 eligible patients in the study population.

### Monthly Rates of Screening and Diagnostic Imaging

We used MGB electronic health record data to determine whether, and when, each participant completed a breast cancer screening procedure and/or diagnostic imaging. We differentiated whether a test was screening or diagnostic using Epic Radiant exam codes and descriptions from radiology reports where ‘SCRN’ identified screening exams and ‘DIAG’ identified diagnostic exams.

The primary outcome of interest was the monthly breast cancer screening rate. This was calculated by dividing the number of patients who received breast cancer screening in each calendar month by the number of patients who were eligible for breast cancer screening in that month. Eligibility for screening during the study period (January 1, 2019 – December 31, 2020) was determined monthly by assessing whether a patient was 40–74 years of age without a documented mammogram within the previous year plus one month (N=2,150). While the U.S. Preventive Services Task Force (USPSTF) recommends biennial mammography screening for women aged 40 to 74 and this recommended interval may vary further across individuals due to prior abnormalities, high-risk status, or healthcare provider or patient decision making,^21^ we used a 1-year period as the expected interval as it was the prevailing screening pattern prior to COVID-19 in this study population. We also excluded participants who were age 75 or older at the start of the study.

The secondary outcome was the monthly rate of breast cancer diagnostic imaging. This was similarly calculated by dividing the counts of breast cancer diagnostic imaging in a calendar month by all eligible participants within the BMCS cohort. Everyone in the study population was eligible for diagnostic imaging, without any age restrictions or prior completed visits necessary (N=2,392).

### Statistical Analysis

We summarized descriptive statistics for eligible study participants within BMCS, including characteristics on age (≥50 years, <50 years), race (White, non-White), and first-degree family history of breast cancer (yes or no). Age was dichotomized at age 50 due to the difference in the risk of developing breast cancer between the two age groups^22,23^. Race was dichotomized due to small numbers of individuals (and screening or diagnostic imaging events) in non-White racial group.

The breast cancer screening rates and diagnostic imaging rates among eligible months for each patient, along with their 95% exact Poisson confidence intervals, were calculated for all patients in the pre-pandemic (January 2019-February 2020), lockdown (March-May 2020), and re-opening (June-December 2020) periods.^24^ This analysis was additionally stratified by age, race, and first-degree family history of breast cancer.

We conducted interrupted time series analyses in which we fit segmented Poisson regression models including covariates for months since January 2019, indicators for lockdown and re-opening periods, and interactions between the time trend and indicators, along with an offset for number of eligible patients in each month.^25,26^ The incidence rate ratio (IRR) for monthly trends in each period and the ratio of these IRRs between periods were estimated based on the model. We also fit expanded models that additionally included indicators for the subgroup, two-way interactions between the time trend and both indicators for period and subgroup, and three-way interactions with both the time trend and indicators to estimate the ratio of time trend IRRs across patient groups in each period.^27^ Separate sets of models were fit for rates of screening and diagnostic procedures. Two-sided p-values were calculated based on Wald tests for differences in trends between the periods and patient groups within periods.

Throughout we used heteroscedasticity and autocorrelation consistent standard errors to account for overdispersion and residual autocorrelation^28,29^. All data analysis was conducted using R statistical software (version 4.1.1).

## RESULTS

Among participants eligible for screening analyses, the mean age was 50.7 (± 7.8) years with 1577 (73.3%) White participants, and 470 (21.9%) participants reporting a first-degree family history of breast cancer (Table 1). Among those who were eligible for the diagnostic imaging analyses, the mean age was 53.0 (± 10.1) years. 1,774 (74.2%) participants were White and 525 (21.9%) reported a first-degree family history of breast cancer.

**Table 1.** Boston Mammography Cohort Study Demographics (N=2392)*

| Table 1. Boston Mammography Cohort Study Demographics (N=2392)* |  |  |  |  |
| --- | --- | --- | --- | --- |
|  | Total Eligible for Screening<br>(N=2150)** |  | Total Eligible for Diagnostic<br>Imaging (N=2392) |  |
| Age*** (years) | 50.7 ± 7.8 |  | 53.0 ± 10.1 |  |
| Race |  |  |  |  |
| White | 1577 (73.3%) |  | 1774 (74.2%) |  |
| Non-White | Total | 573 (26.7%) | Total | 618 (25.8%) |
|  | Black | 252 (44.0%) | Black | 280 (45.3%) |
|  | Native American | 11 (1.9%) | Native American | 13 (2.1%) |
|  | Asian | 54 (9.4%) | Asian | 57 (9.2%) |
|  | Hawaiian | 2 (0.3%) | Hawaiian | 2 (0.3%) |
|  | Unknown | 242 (42.2%) | Unknown | 254 (41.1%) |
|  | More than one race | 12 (2.1%) | More than one race | 12 (2.0%) |
| Family History |  |  |  |  |
| Yes | 470 (21.9%) |  | 525 (21.9%) |  |
| No | 1680 (78.1%) |  | 1867 (78.1%) |  |
| *Reported N (%) or Mean ± SD |  |  |  |  |
| ** Additional eligibility criteria for screening: 40–74 years of age without a documented mammogram in the previous year. |  |  |  |  |
| *** Age at study baseline. |  |  |  |  |

As shown in Table 2, from January 2019 to December 2020, the breast cancer screening rate in the BMCS cohort overall was 45.1 per 1,000 person-months during the pre-COVID-19 period, 7.0 per 1,000 person-months during the lockdown period, and 49.6 per 1,000 person-months during the reopening period. As for breast cancer diagnostic imaging rate, it was 5.7 per 1,000 person-months during the pre-COVID-19 period, 2.9 per 1,000 person-months during the lockdown period, and 6.3 per 1,000 person-months during the reopening period. This is also reflected in Figure 1 where the initial sharp decline in both the screening and diagnostic imaging rates after March 2020 during the lockdown period was then followed by a rebound in the rates during the reopening period.

**Table 2.** Rates (per 1,000 person-months) of breast cancer screening and diagnostic imaging before COVID-19, during lockdown, and during the reopening period, overall and by age, race, and family history*

|  |  |  |  |
| --- | --- | --- | --- |
|  | **Incidence Rate (95% Confidence Interval)** |  |  |
|  | Pre-COVID-19 | Lockdown | Reopening |
| **Screening** |  |  |  |
| Overall | 45.1 (42.2, 48.1) | 7.0 (4.6, 9.9) | 49.6 (45.5, 53.9) |
| Age |  |  |  |
| Younger than 50 | 31.5 (24.9, 39.5) | 4.6 (0.6, 16.7) | 33.9 (23.2, 47.9) |
| Age 50 and older | 46.8 (43.8, 50.1) | 7.3 (4.9, 10.4) | 51.0 (46.8, 55.6) |
| Race |  |  |  |
| Non-White | 34.0 (29.6, 39.0) | 7.8 (3.7, 14.4) | 34.9 (28.6, 42.0) |
| White | 49.6 (46.1, 53.4) | 6.7 (4.1, 10.2) | 55.4 (50.4, 60.8) |
| Family History |  |  |  |
| Yes | 44.0 (40.9, 47.3) | 6.0 (3.7, 9.1) | 49.1 (44.6, 54.0) |
| No | 49.2 (42.8, 56.3) | 11.0 (5.3, 20.2) | 51.5 (42.6, 61.7) |
| **Diagnostic** |  |  |  |
| Overall | 5.7 (4.9, 6.6) | 2.9 (1.8, 4.5) | 6.3 (5.2, 7.7) |
| Age |  |  |  |
| Younger than 50 | 5.7 (4.1, 7.7) | 2.0 (0.4, 5.8) | 5.7 (3.5, 8.8) |
| Age 50 and older | 5.7 (4.8, 6.7) | 3.2 (1.9, 5.0) | 6.5 (5.2, 8.0) |
| Race |  |  |  |
| Non-White | 5.9 (4.4, 7.8) | 2.2 (0.6, 5.5) | 5.8 (3.7, 8.5) |
| White | 5.6 (4.7, 6.7) | 3.2 (1.9, 5.1) | 6.5 (5.2, 8.1) |
| Family History |  |  |  |
| No | 5.7 (4.8, 6.7) | 1.8 (0.9, 3.3) | 6.6 (5.3, 8.1) |
| Yes | 5.7 (4.1, 7.7) | 7.0 (3.5, 12.5) | 5.4 (3.3, 8.4) |
| * Pre-COVID-19 is defined as the time frame from Jan 1, 2019 – Feb 29, 2020; the lockdown period is defined as the time frame from Mar 1 – May 30, 2020; the reopening period is defined as the time frame from Jun 1 – Dec 31, 2020. |  |  |  |

**Figure 1.**
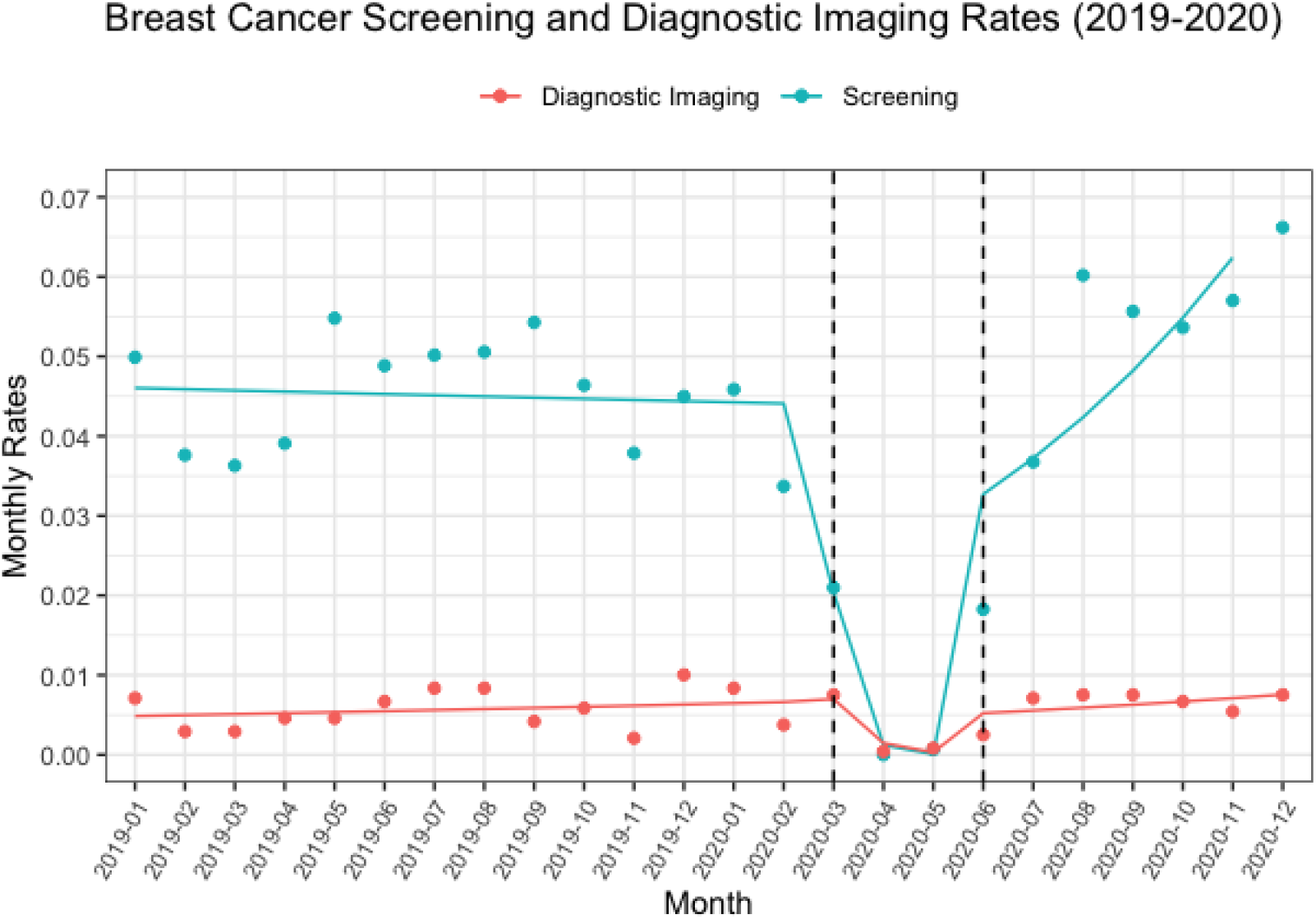

Comparing the overall rates across the three time periods, we observed a 94% decline in the monthly trend of breast cancer screening rates (p<0.001) and a 79% decline in the monthly trend of diagnostic imaging rates (p=0.002) during the lockdown period compared to the pre-COVID-19 period (Table 3). During the reopening period, we observed a 14% increase in the monthly trend of breast cancer screening rates (p=0.005) compared to the pre-COVID-19 period, but no statistically significant difference in the monthly trend of diagnostic imaging rates compared to the pre-COVID-19 period (p=0.580).

**Table 3.** The monthly trend and incidence rate ratio of expected monthly rate of breast cancer screening and diagnostic imaging overall *

| Time Periods | Screening |  |  | Diagnostic |  |  |
| --- | --- | --- | --- | --- | --- | --- |
|  | Monthly Trend IRR (95% CI) | Ratio of IRRs** | p-value | Monthly Trend IRR (95% CI) | Ratio of IRRs** | p-value |
| **Pre-COVID-19** | 0.997<br>(0.981, 1.013) | Reference | Reference | 1.024<br>(0.989, 1.061) | Reference | Reference |
| **Lockdown** | 0.060<br>(0.015, 0.231) | 0.06 | <0.001 | 0.205<br>(0.088, 0.482) | 0.21 | 0.002 |
| **Reopening** | 1.142<br>(1.091, 1.195) | 1.14 | 0.005 | 1.039<br>(0.938, 1.150) | 1.04 | 0.580 |

\* Pre-COVID-19 is defined as the time frame from Jan 1, 2019 – Feb 29, 2020; the lockdown period is defined as the time frame from Mar 1 – May 30, 2020; the reopening period is defined as the time frame from Jun 1 – Dec 31, 2020.
\*\* Incidence rate ratios (IRR) and ratio of IRRs estimated from an interrupted time series analysis with Poisson regression and robust standard errors.

During the pre-COVID-19 period, the monthly trend of breast screening was 5% higher among women aged 50 and older compared to women younger than 50 (p=0.005) (Table 4, Figure 2A). During the same period, women aged 50 and older had a 10% higher monthly trend of breast cancer diagnostic imaging rates (p=0.039) compared to women younger than 50 (Table 4, Figure 2A). There was no statistically significant difference in either the monthly trend of screening or diagnostic imaging rates between White and non-White women, or by family history status during the pre-COVID-19 period.

**Table 4.** The monthly trend and incidence rate ratio of expected monthly rate of breast cancer screening and diagnostic imaging overall and by age, race, and family history *

|  |  |  |  |  |  |  |  |
| --- | --- | --- | --- | --- | --- | --- | --- |
|  |  | Screening |  |  | Diagnostic |  |  |
|  |  | Monthly Trend IRR | Ratio of IRRs** | p-value | Monthly Trend IRR | Ratio of IRRs** | p-value |
| The Pre-COVID-19 Period |  |  |  |  |  |  |  |
| Age | Younger than 50 | 0.950 | Ref | 0.005 | 0.952 | Ref | 0.039 |
|  | 50 and older | 1.000 | 1.05 |  | 1.045 | 1.10 |  |
| Race | Non-White | 0.997 | Ref | 0.987 | 1.023 | Ref | 0.976 |
|  | White | 0.997 | 1.00 |  | 1.025 | 1.00 |  |
| Family History | No | 0.997 | Ref | 0.918 | 1.016 | Ref | 0.163 |
|  | Yes | 0.995 | 1.00 |  | 1.053 | 1.04 |  |
| The Lockdown Period |  |  |  |  |  |  |  |
| Age | Younger than 50 | 1.010 | Ref | <0.001 | 0.594 | Ref | 0.037 |
|  | 50 and older | 0.000 | 0.00 |  | 0.150 | 0.25 |  |
| Race | Non-White | 0.000 | Ref | <0.001 | 0.000 | Ref | <0.001 |
|  | White | 0.086 | NC*** |  | 0.257 | NC*** |  |
| Family History | No | 0.000 | Ref | <0.001 | 0.262 | Ref | 0.723 |
|  | Yes | 0.175 | NC*** |  | 0.163 | 0.62 |  |
| The Reopening Period |  |  |  |  |  |  |  |
| Age | Younger than 50 | 1.381 | Ref | 0.001 | 0.904 | Ref | 0.190 |
|  | 50 and older | 1.125 | 0.81 |  | 1.105 | 1.22 |  |
| Race | Non-White | 1.122 | Ref | 0.805 | 0.840 | Ref | 0.018 |
|  | White | 1.142 | 1.02 |  | 1.144 | 1.36 |  |
| Family History | No | 1.144 | Ref | 0.741 | 1.051 | Ref | 0.481 |
|  | Yes | 1.114 | 0.97 |  | 1.120 | 1.07 |  |
| * Pre-COVID-19 is defined as the time frame from Jan 1, 2019 – Feb 29, 2020; the lockdown period is defined as the time frame from Mar 1 – May 30, 2020; the reopening period is defined as the time frame from Jun 1 – Dec 31, 2020. |  |  |  |  |  |  |  |
| ** Incidence rate ratios (IRR) and ratio of IRRs estimated from an interrupted time series analysis with Poisson regression and robust standard errors. |  |  |  |  |  |  |  |
| ***NC: This IRR is not calculated (NC) and not reported because the IRR for the reference group is extremely low (approaching zero) resulting in extremely large ratio of IRRs (approaching infinity) that are not meaningfully interpretable. |  |  |  |  |  |  |  |

**Figure 2.**
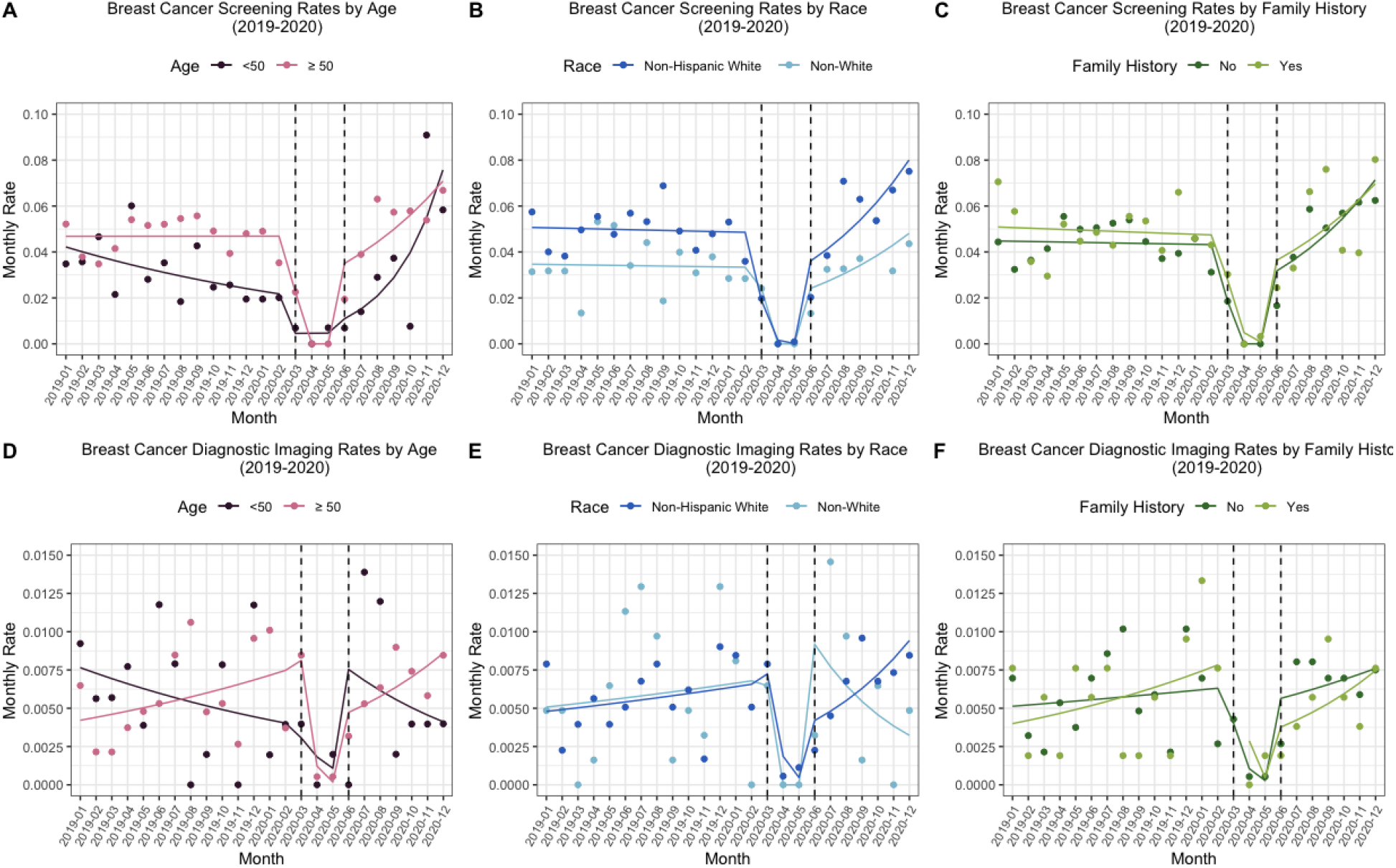

During the lockdown period (Table 4), women aged 50 or older had approximately 100% lower monthly trend of breast cancer screening rates (p<0.0001) and a 75% lower monthly trend of breast cancer diagnostic imaging rates (p=0.037) compared to that of women younger than 50. In fact, women aged 50 or older had no breast cancer screenings during this period. In the same period, those who were White had higher monthly trends of both breast cancer screening and diagnostic imaging rates compared to those who were non-White (p<0.0001) (Figure 2B & 2E). Those with a family history of breast cancer in a first degree relative also had a higher monthly trend of breast cancer screening rates compared to those without family history (p<0.0001), while the observed 38% decrease in the monthly trend of breast cancer diagnostic rates did not differ between those with or without family history (p=0.723) (Figure 2C & 2F). Despite these differences, the monthly trends of both breast cancer screening and diagnostic imaging rates were extremely low for all women during the lockdown period, regardless of race and family history (p<0.0001) (Figure 2B, 2C, 2E, 2F).

During the reopening period (Table 4), women aged 50 or older had a 19% lower monthly trend in the screening rate compared to that among women younger than 50 (p=0.001) and those who were White had a 36% higher monthly trend in the breast cancer diagnostic imaging rate in comparison to those who were non-White (p=0.018). There was no significant difference in the monthly trend of screening rates between White and non-White women, or a significant difference in diagnostic imaging rates between women younger than 50 and women aged 50 and older. We also did not observe a significant difference in either the monthly trend of screening or the diagnostic imaging rates according to family history during the reopening period.

## DISCUSSION

We observed a decline in overall breast cancer screening and diagnostic imaging rates with the advent of the COVID-19 pandemic. Screening and diagnostic imaging rates rebounded during the reopening phase, equaling, or exceeding those of the pre-COVID-19 period, which may reflect additional screens performed to address the backlog of missed visits during the lockdown. These results for the overall trends for breast cancer screening and diagnostic imaging rates are consistent with findings from prior studies^11,18,19^. Furthermore, we observed some variation in breast cancer screening and diagnostic imaging monthly trends by age and race. During the pre-COVID-19 period, the monthly trend of breast cancer screening rates was higher among women aged 50 or older compared to those younger 50, but this reversed during the reopening phase with higher monthly trends among women younger than 50 in comparison to women aged 50 or older (Table 4). While we observed no difference in the monthly trend of diagnostic imaging rates by race prior to or during the lockdown, during the reopening phase imaging rates were trending down for non-White women and trending up for White women.

Interestingly, we observed no variation in the monthly trends of breast cancer screening or diagnostic imaging rates by family history of breast cancer pre-COVID-19 or during the recovery period. However, this may also be explained by low power due to few events among the small number of women with a family history of breast cancer. Considering that women aged 50 or older are already at higher risk of breast cancer and non-White women tend to have more advanced stage of disease at the time of diagnosis and are more likely to die from breast cancer^30–33^, a slower re-uptake of breast cancer diagnostic visits among non-White women warrants concerns for worsened disparities in breast cancer outcomes due to the COVID-19 pandemic. Careful attention must be paid to these vulnerable subgroups as the COVID-19 recovery continues to ensure equitable resumption of care.

Our findings are consistent with prior studies of COVID-19 related changes in breast cancer screening and diagnostic imaging and potential disparities during this period. Nyante et al. found an 85.1% reduction for screening mammography and a 48.9% reduction in diagnostic mammography in March 2020 immediately after the pandemic’s onset^11^. Similarly, Velazquez et al. compared the proportion of completed mammograms out of the total scheduled between pre- and post-COVID-19 and found a 17% decrease, where the proportion of completed mammograms among Black women was the lowest^19^. Additionally, Marcondes et al. assessed changes in rates of breast cancer screening by race within the Mass General Brigham healthcare system pre- and post-COVID-19 and found that Latinx individuals had significantly lower rates of breast cancer screening post-COVID-19 in comparison to pre-COVID-19.

However, the authors found no significant worsening or improvement of racial/ethnic disparities for breast cancer screening, which is consistent with our study results. They did not examine breast related diagnostic imaging rates or examine rates by age group or family history of breast cancer^18^. In addition, Sprague et al. found that changes in breast cancer screening mammography during the pandemic were similar across women with and without family history of breast cancer among the Breast Cancer Surveillance Consortium^34^. While Labaki et al. did not examine subgroup differences, they did find modest age variation among patients undergoing mammography between different time periods pre- and post-COVID-19, but that was only when they used the three immediate months prior to the lockdown orders as their reference group^35^.

Differences in trends of diagnostic imaging incidence between White and non-White women during the reopening period could be driven by differences in abnormal findings from screening exams or differences in presentation of symptoms or some combination. However, as there were no racial differences in screening during the shutdown or reopening periods, it suggests that our findings were due to racial differences in referrals, follow-up with diagnostic imaging after screening, or recognizing and receiving care for breast symptoms during the reopening period. These results are in line with previous research that has found delays from screening to diagnostic imaging. A study conducted by Miller-Kleinhenz et al., assessed factors that contribute to the delay from screening to diagnosis, including diagnostic imaging in a cohort of women diagnosed with breast cancer in the metropolitan Atlanta hospital system. Results from this study demonstrate that systemic inequities such as provider biases, discrimination, social and physical environments may be driving differences in diagnostic imaging.^36^

Our study has several important limitations. First, it was conducted among participants enrolled in a breast cancer mammography cohort at a large academic healthcare system where screening and diagnostic rates may be higher than the general population and may affect generalizability. Second, follow-up for participants did not account for whether patients continued to receive care at MGB as they may have moved prior to the study or during the study period. If participants moved or ceased receiving care in this health system prior to the study, they would not have received screening in any phase of the analysis. Participants that moved because of the pandemic may have received imaging prior to the pandemic, but not in the reopening period, which could affect results if moving differed by family history, age, or race.

Third, we had to combine all non-White individuals into one broad category to have sufficient power for the analysis, which loses the ability to look at potential differences in screening and diagnostic rebound between each racial and ethnic groups. Finally, for interrupted time series analysis comparing across time periods, the analysis with a short series could be subject to confounding by seasonal effects. It is unclear whether such effects could have differential impacts among different age, race, and family history subgroups to bias comparisons.

Our results demonstrate the ripple effects of the COVID-19 pandemic with respect to breast cancer prevention and early detection and portends potential downstream effects on stage at diagnosis and morbidity. Future direction could look at how variation in imaging trends impacts future breast cancer incidence and survival among this cohort. Additionally, future work could examine the rate of breast cancer screening and diagnostic imaging by other demographic characteristics, including insurance status, breast cancer risk scores, and geographic location, and looking across a more extended follow-up period to assess whether health disparities created by COVID-19 persist or alleviate as the pandemic progresses.

## Data Availability

Data used here may be requested through the Boston Mammography Cohort Study research team upon request and approval of the institutional IRB. The data are not publicly available to protect the privacy of research participants.

## DISCLOSURES

### Ethics Statement

The study was approved by the Mass General Brigham Human Research Committee, the Institutional Review Board (IRB) of Mass General Brigham. Participants provided written informed consent prior to joining the Boston Mammography Cohort Study.

